# Fall Frequency, Risk Factors, and Outcomes in Parkinson’s Disease: A Cross-Sectional Analysis

**DOI:** 10.1101/2025.08.05.25332959

**Authors:** Joaquin A. Vizcarra, Kat Hefter, David-Erick Lafontant, Michael Tran Duong, Ashkan Ertefaie, Brian Litt, Dani S. Bassett, Andrew Siderowf, The Parkinson’s Progression Markers Initiative

**Affiliations:** Department of Neurology, University of Pennsylvania Perelman School of Medicine, Philadelphia, PA, USA; Center for Neuroengineering and Therapeutics, University of Pennsylvania, Philadelphia, PA 19104, USA; Department of Bioengineering, University of Pennsylvania School of Engineering and Applied Sciences, Philadelphia, PA, USA; Department of Biostatistics, College of Public Health, University of Iowa, Iowa City, IA, USA; Department of Radiology, University of Pennsylvania Perelman School of Medicine, Philadelphia, PA, USA; Department of Biostatistics, Epidemiology & Informatics, University of Pennsylvania, Philadelphia, PA, USA

**Author notes:** Corresponding Author: Joaquin A. Vizcarra, MD Department of Neurology, University of Pennsylvania Perelman School of Medicine 330 S 9^th^ St, Philadelphia, PA, USA 19107. These two authors contributed equally to this work (Joaquin A. Vizcarra, Kat Hefter).

**Keywords:** Parkinson, fall, NSD-ISS, risk factors

## Abstract

**Background:** Falls are a major source of morbidity in Parkinson’s disease (PD), yet their evolution over time remains unclear.

**Objectives:** To compare fall risk and outcomes among PD, prodromal alpha-synucleinopathy, (PAS) and healthy controls (HC); estimate fall frequency across PD progression; and characterize clinical features of PD faller subgroups.

**Methods:** We analyzed fall-related outcomes in the Parkinson’s Progression Markers Initiative. Yearly rates of rare and frequent falls were estimated by time since diagnosis. Unique PD participants were grouped as never, rare, or frequent fallers. Clinical variables included motor, cognitive, behavioral, sleep, and autonomic measures. Outcomes included injuries and healthcare utilization. Regression models adjusted for age, sex, and disease duration, with Benjamini-Hochberg correction.

**Results:** Across 6,977 visits from 3,100 participants (937 PD, 1,926 PAS, 237 HC), PD participants had higher odds of falling than PAS (OR=1.66, 95% CI [1.46–1.87]) and HC (OR=4.03, 95% CI [3.14–5.23]). PD participants were also more likely to report fall-related injuries and healthcare use than PAS (OR=1.70, 1.71) and HC (OR=3.26, 3.81). Falls occurred in 15.5% of visits at diagnosis and 69.2% after 14 years, increasing across Neuronal Synuclein Disease-Integrated Staging System (NSD-ISS) stages. Frequent fallers had longer disease duration, higher NSD-ISS, and worse clinical profiles. Women were more likely to fall than men (46.1% vs 34.9%, p=0.002) despite milder symptoms.

**Conclusion:** Falls and related morbidity increase with disease duration and NSD-ISS. Risk reflects sex and motor and non-motor factors, supporting a multifactorial model. Fall frequency may represent a practical marker of progression and guide prevention strategies in PD.

## INTRODUCTION

Falls are a common and important source of morbidity and mortality in people with Parkinson’s disease (PwP). PwP fall twice as often as age-matched individuals without Parkinson’s disease (PD),^1^ with 60% experiencing at least one fall,^2^ and 9% transitioning annually from no or rare to monthly falls.^3^ Consequences include fractures, hospitalizations, and reduced quality of life.^4^ PwP have a three-fold increased risk of hip fracture,^5^ and post-fracture mortality is twice than those without PD.^6^ A substantial share of the $7.1 billion USD in excess PD-related healthcare costs in 2017 is likely due to falls,^7^ though specific costs remain unquantified.

Several clinical risk factors have been identified. Prior falls are among the strongest predictors.^8^ Motor impairment, such as freezing of gait, postural instability, and higher Hoehn & Yahr (H&Y) and Movement Disorders Society Unified Parkinson’s Disease Rating Scale (MDS-UPDRS) Part III scores are also associated with fall risk.^9^ The postural instability/gait difficulty (PIGD) phenotype carries greater risk than tremor-dominant PD.^2^ Cognitive impairment, including global decline, slowed processing speed,^10,11^ and domain-specific inattention and executive dysfunction,^12^ also contribute. Autonomic dysfunction, particularly neurogenic orthostatic hypotension (OH), has been associated with falls due to fainting.^13^ Importantly, recent work on PD disease milestones suggests that many of these risk factors cluster as the disease advances, with motor, cognitive, and autonomic impairments co-occurring and compounding fall risk.^14^ However, current knowledge largely derives from retrospective or short-duration studies, limiting insights into fall evolution over time.

Using the Parkinson’s Progression Markers Initiative (PPMI) dataset, we aimed to: (1) compare the risk of falls and outcomes among PD, prodromal alpha-synucleinopathy (PAS), and healthy controls (HC); (2) estimate fall frequency across PD duration and biological stages; and (3) characterize fall-related clinical features and outcomes in fall-prone PD patients. Our goal is to identify at-risk patients to inform integrated prevention strategies.

## METHODS

### Standard Protocol Approvals, Registrations, and Patient Consents

The study was approved by institutional review boards at all PPMI sites, and participants provided written informed consent. Data, protocol, and materials may be requested from the PPMI at https://www.ppmi-info.org. This cross-sectional study followed STROBE reporting guidelines.^15^

### Study design

PPMI is a multicenter, international, prospective cohort study.^16^ We first assessed fall frequency and outcomes at each visit among PD, PAS, and HC participants. We then created a cross-sectional dataset from PD visits to form adequately sized groups of unique PwP with different fall frequencies. Each PwP was included once and assigned to a single group, as detailed in the statistical analysis.

### Participants

Briefly, PwP were required to be aged 30 years or older (at diagnosis); have a H&Y score of <3; have at least two of the following: resting tremor, bradykinesia, rigidity (must have either resting tremor or bradykinesia), OR either asymmetric resting tremor or asymmetric bradykinesia; and evidence of dopaminergic deficit consistent with PD based on dopamine transporter single-photon emission computed tomography (DAT SPECT) imaging. PAS participants had hyposmia and/or REM Sleep Behavior Disorder (RBD) with mild deficits on DAT SPECT, without pathogenic genetic variants that increase the risk of developing PD. HC had no clinically significant neurological disorder, no first-degree relative with PD, and normal DAT SPECT.

Data sources included: 1. Curated public data cut (v.20241211) and 2. Determination of Freezing and Falls (v.20250110). We excluded participants with known monogenic PD and those without any fall data.

### Clinical variables

PPMI participants had comprehensive evaluations at annual study visits.^17^ We analyzed demographic and clinical variables, including age, sex (assigned at birth), race, ethnicity, years of education, body mass index (kg/m2), disease duration (months from diagnosis), levodopa equivalent daily dose (LEDD),^18^ modified Schwab and England Activities of Daily Living Scale (S&E), and Neuronal Synuclein Disease Integrated Staging System (NSD-ISS).^19^ We also analyzed assessments of motor, cognitive, behavioral, sleep, and autonomic measures.^14^ Motor assessments included MDS-UPDRS, H&Y stage, and PIGD scores.^20^ Cognitive assessments included Montreal Cognitive Assessment (MoCA), Benton Judgment of Line Orientation (BJLO), Clock Drawing Test, Modified Boston Naming Test (MBNT), Hopkins Verbal Learning Test (HVLT), Symbol Digit Modalities Test (SDMT), and Trails Making Test (TMT). Behavioral assessments included Questionnaire for Impulsive-Compulsive Disorders in Parkinson’s Disease (QUIP), Geriatric Depression Scale (GDS-15), and State-Trait Anxiety Inventory (STAI). Sleep assessments included the Epworth Sleepiness Scale (ESS) and RBD Screening Questionnaire (RBDSQ). Autonomic assessments included Scales for Outcomes in Parkinson’s disease – Autonomic Dysfunction (SCOPA) and orthostatic (supine to standing) change in systolic blood pressure. No data imputation was performed for missing clinical variables.

### Determination of falls, injury, and healthcare utilization

We assessed falls using self-reported modified items 13 and 14 of the Unified Parkinson’s Disease Rating Scale (UPDRS) – Part II,^21^ introduced in PPMI in 2019 for yearly collection. Item 13 describes falls unrelated to freezing of gait, whereas item 14 describes falls related to freezing, within the last week and 12 months. We combined responses in both time frames and selected the one with higher scores. Fall frequency was defined as “none”, “rare”, and “frequent” based on combinations reported in eTable 1. We assessed fall-related injury and healthcare utilization via self-reported binary questionnaires about the different fall-related injuries and healthcare utilization in the past year. We collected the following injuries: hip fracture, upper extremity fracture, skull fracture, other fracture, head injury, laceration requiring sutures, and other injuries. We collected data on the healthcare utilizations: outpatient clinic visit, emergency department visit, inpatient hospitalization, surgery, and institutionalization. We determined fall frequency, injury, and healthcare utilization at yearly visits. No data was imputed when fall frequency was not recorded. For visits with documented falls but missing data on injury or healthcare utilization, these outcomes were assumed not to have occurred. In all other cases, missing injury or healthcare utilization data were left blank.

### Statistical analysis

All statistical analyses were conducted using R 4.5.0 (R Foundation for Statistical Computing, Vienna, Austria).

#### Frequency of falls and fall-related outcomes between PD, PAS, and HC

We analyzed falls, injuries, and healthcare utilization across all cohorts and years. Logistic regression models, with PD as the reference, estimated outcome likelihood adjusting for age and sex. Wald tests assessed cohort effects; when significant (i.e., cohort membership was significant), odds ratios (ORs) were derived for PD versus other cohorts and females versus males. Multiple comparisons were corrected with the Benjamini-Hochberg (BH) procedure (False discovery rate [FDR] = 0.05, p<0.028). We then evaluated fall frequency effects on injuries and healthcare utilization in PD and PAS cohorts using logistic models with age, sex, and cohort as covariates. BH correction was again applied (FDR = 0.05, p<0.024). ORs were calculated for PD/PAS, frequent/rare fallers, and female/male participants.

#### Frequency of falls and fall-related outcomes across disease duration in PD

In the PD cohort, fall frequency was assessed yearly and categorized by disease duration (rounded to the nearest year) and NSD-ISS stage. We calculated the proportion of visits with falls, injuries, and healthcare utilization for each duration year and stage. Individual PwP could contribute data to multiple years if fall data were available.

#### Correlates of falling between faller groups and sex in PD

Because the fall questionnaire was introduced in 2019, earlier enrollees often lacked fall data in the initial years, creating incomplete longitudinal records. To reduce confounding from unmodeled repeated measures and improve reliability of classification, we required at least two consecutive years of fall data for sampling. This approach limited bias from single-year variability or reporting error, while ensuring consistent group assignment across participants.

Each individual contributed a single observation and was classified into one of three mutually exclusive groups: never, rare, or frequent fallers. Participants’ first occurrence of either “rare-frequent” or “frequent-frequent” fall years were labeled as “frequent fallers”, and available variables from the last observation in that timeframe were taken. These individuals were then removed from the pool of participants who could be considered as either “never fallers” or “rare fallers.” Then, participants’ first occurrence of the combination of either “none-rare” or “rare-rare” fall years were labeled as “rare fallers,” available variables from the last observation in that timeframe were taken, and those participants were removed from the pool of participants who could be considered as “never fallers.” Finally, participants’ first occurrence of two consecutive “none” years were sampled and labeled as “never fallers,” and available variables from the last observation in that time frame were taken. Based on this process, an individual subject could only be counted in one category, even if they progressed from being a never faller to a frequent faller over the subsequent study visits. Due to low event counts, injuries or healthcare use were recorded if reported in either of the two sampled years.

We then compared fall groups and sexes. We summarized variables using counts and proportions, as well as means and standard deviations, as appropriate. For categorical and continuous variables, we used logistic models and linear regression models, respectively, with age, sex, and years since diagnosis as covariates. For pairwise comparisons between fall frequency groups, the reference group was set to “rare fallers.” We used BH correction for multiple comparisons (FDR = 0.05, p<0.016).

## RESULTS

Across cohorts, 3,805 individuals were evaluated for inclusion: 1,442 with PD, 2,045 PAS, and 318 HC. We excluded 360 monogenic PD patients and 705 participants without fall data (e.g., 505 PD, 119 PAS, 81 HC), yielding a final sample of 3,100 (e.g., 937 PD, 1,926 PAS, 237 HC). In total, 6,977 visits were analyzed (e.g., 2,372 PD, 3,848 PAS, 757 HC). Mean ages across visits with fall data were 66.8 y (SD ± 9.41) for PD, 68.1 y (SD ± 6.54) for PAS, and 67.9 y (SD ± 11.5) for HC. The proportion of male participants was highest in the PD group (66.2%), followed by HC (61.0%) and PAS (48.8%). Falls were reported in 1,607 visits. In PD, 514 visits had rare falls, and 163 visits had frequent falls (21.7% and 6.9%, respectively); in PAS, 806 visits had rare falls, and 44 visits had frequent falls (20.9% and 1.1%, respectively); and in HC, 78 visits had rare falls, and two visits had frequent falls (10.3% and 0.3%, respectively).

### Comparison of risk of falls, injury, and healthcare utilization between PD, PAS, and HC

In the sample of 3,100 participants across 6,977 visits, PwP were more likely to report falls, fall-related injury, and healthcare utilization than PAS and HC (eTable 2). Adjusted analyses showed PwP had 1.66 times the odds of falling versus PAS (95% CI: 1.46, 1.87), and 4.03 times the odds versus HC (95% CI: 3.14, 5.23). Odds of injury were 1.70 versus PAS (95% CI: 1.42, 2.04), and 3.26 versus HC (95% CI: 2.27, 4.85). Odds of healthcare utilization were 1.71 versus PAS (95% CI: 1.39, 2.09), and 3.81 versus HC (95% CI: 2.48, 6.12). When fall frequency was added to the models, cohort membership no longer predicted injury or healthcare utilization (eTable 3), whereas fall frequency remained a robust independent predictor (eTable 4).

### Falls, injury, and healthcare utilization in PD across years since diagnosis and NSD-ISS stages

In 937 participants with PD across 2,372 visits, fall frequency increased with disease progression and NSD-ISS stages. At 0, 7, and 14 years of years since diagnosis, falls occurred in 15.5%, 36.3%, and 69.2% of visits, respectively (Figure 1A). At those same time points, rare falls were reported in 14.0%, 25.3%, and 38.5% of visits, while frequent falls were reported in 1.4%, 11.0%, and 30.8% of visits, respectively. Injury and healthcare utilization also increased with years since diagnosis (Figure 1B). At 0, 7, and 14 years of years since diagnosis, fall-related injuries were reported in 4.3%, 14.3%, and 46.2% of visits, and fall-related healthcare utilization was reported in 2.5%, 9.9%, and 30.8% of visits, respectively. Among visits with falls, the proportion with a fall-related injury increased with years since diagnosis, reaching 27.9%, 39.4%, and 66.7% at years 0, 7, and 14, respectively.

**Figure 1:**
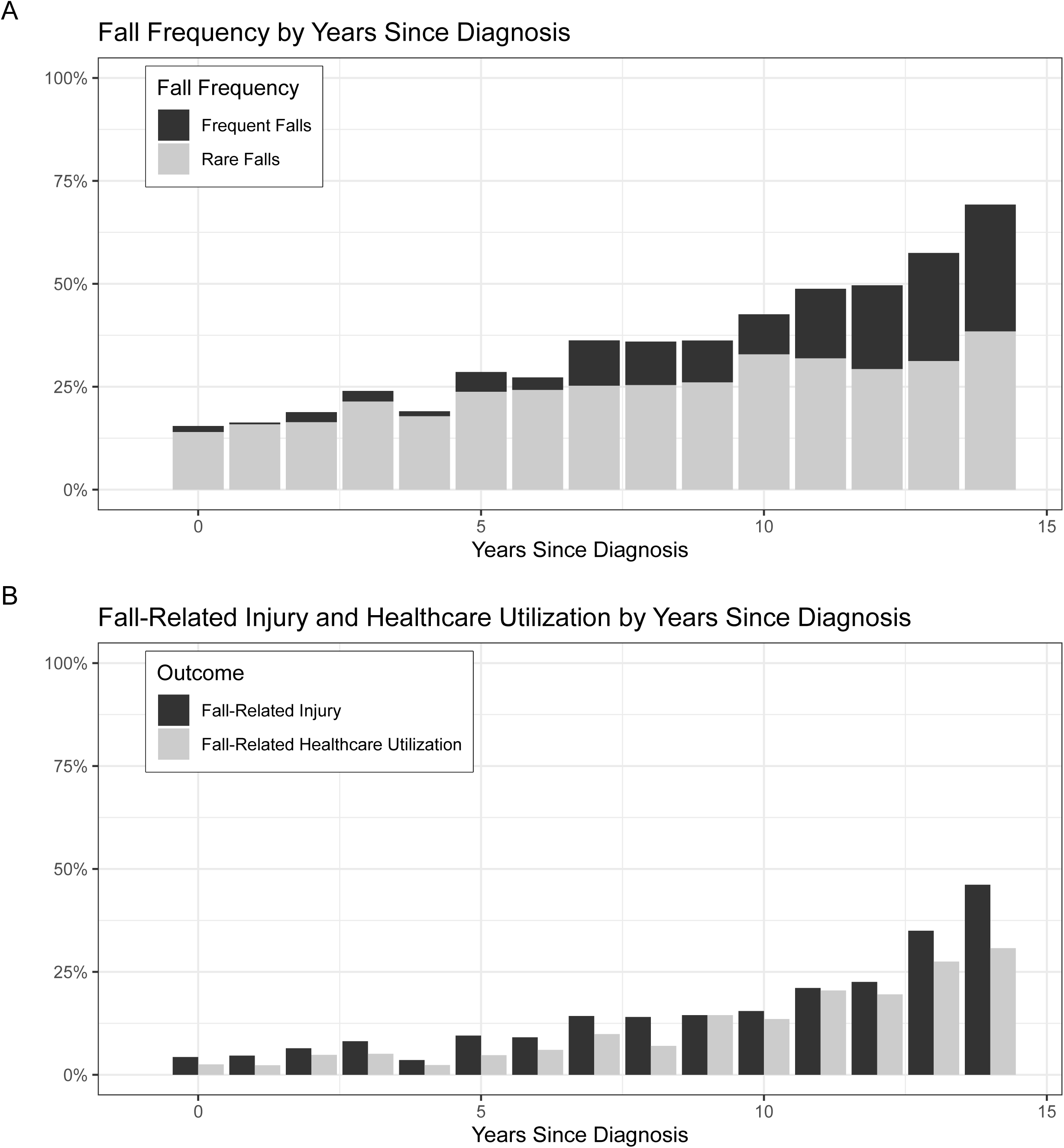
**Fall Frequency and Outcomes across Years Since Diagnosis in PD** In 937 patients with PD across 2372 yearly visits, the percentage of visits with (A) rare or frequent yearly falls and (B) fall-related injury and healthcare utilization is displayed across years since diagnosis.

NSD-ISS data were available for 92.1% of visits (eTable 5). Falls were absent in NSD-ISS stage 2A and limited to rare falls in stage 2B, but increased with higher disability stages. Frequent falls were reported in 2.1% of visits at stage 3, 12.8% at stage 4, 47.0% at stage 5, and 77.8% at stage 6 (Figure 2A). Injuries and healthcare utilization also increased with NSD-ISS stages (Figure 2B). In NSD-ISS stages 3, 4, 5, and 6, injuries were reported in 7.1%, 18.0%, 44.6%, and 77.8% of visits, and healthcare utilization in 5.4%, 14.8%, 34.9%, and 66.7% of visits, respectively.

**Figure 2:**
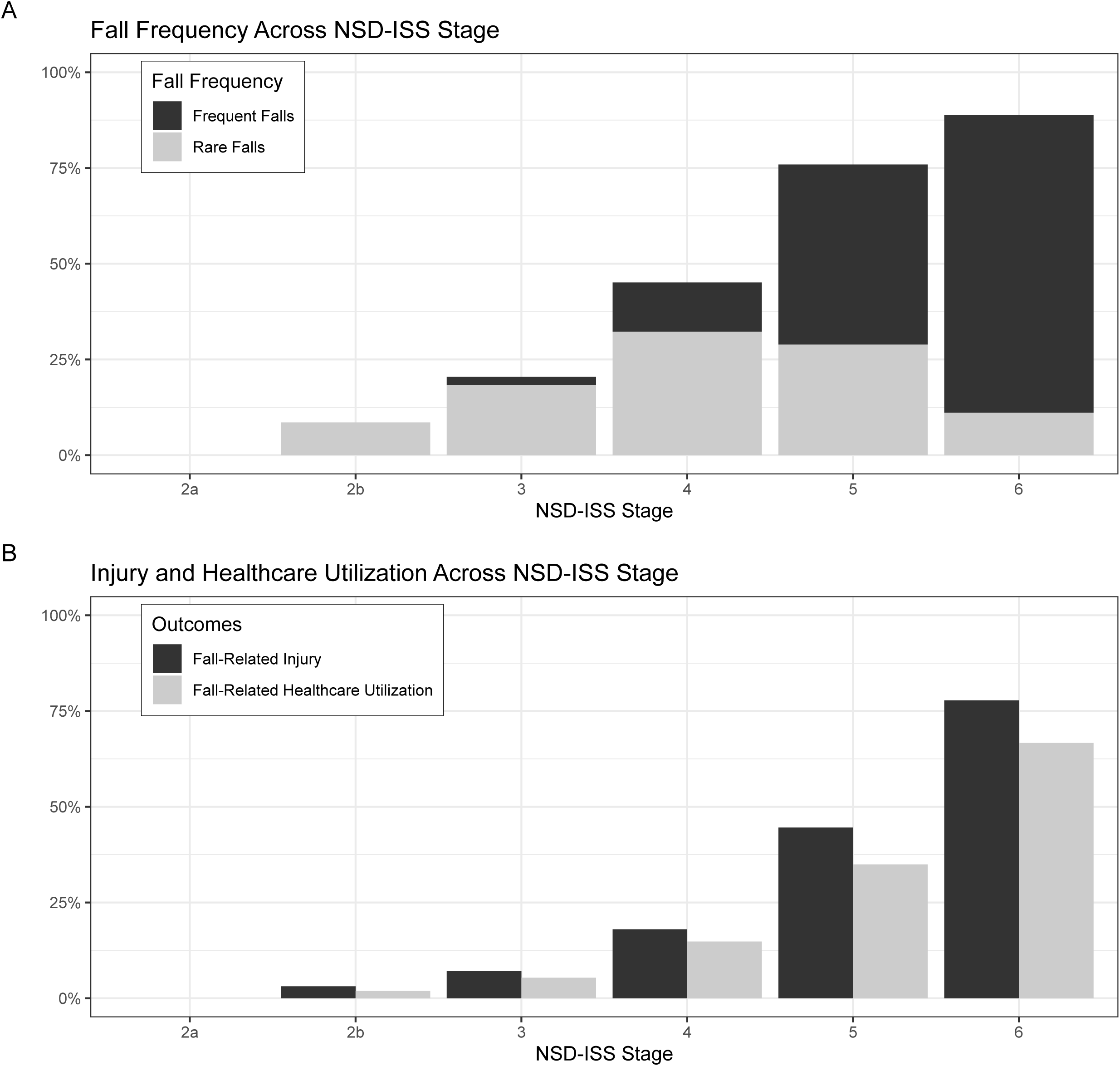
**Fall Frequency and Outcomes across NSD-ISS in PD** Of the 2372 visits, 2185 had NSD-ISS data. A total of 150 visits were of participants labeled not NSD-ISS (i.e., Parkinsonism not from biologically defined neuronal synuclein disease), resulting in 2035 visits with NSD-ISS. The percentage of visits with (A) rare or frequent yearly falls and (B) fall-related injury and healthcare utilization is displayed across NSD-ISS stages. NSD-ISS, Neuronal Synuclein Disease-Integrated Staging System.

### Cross-sectional differences between never, rare, and frequent fallers in unique participants with PD

We classified 371, 178, and 56 unique participants as never, rare, and frequent fallers, respectively (eFigure 1). After adjusting for age and years since diagnosis, male sex was more common in both never and frequent fallers compared to rare fallers (Table 1). Female PwP were significantly more likely than males to report any fall (46.1% versus 34.9%; eTable 6). This difference was driven by rare (39.7% versus 24.2%), but not frequent falls (6.4% versus 10.7%). Years since diagnosis were longest among frequent fallers and shortest among never fallers, with NSD-ISS stage showing a similar gradient. Logistic regression confirmed fall frequency correlated with NSD-ISS stage, with never fallers more often in early stages and frequent fallers in late stages, both relative to rare fallers. S&E scores were lowest in frequent fallers, indicating greater impairment, while LEDD differences were not significant.

**Table 1:** Demographic Differences Between Never, Rare, and Frequent Fallers in the Cross-Sectional PD Cohort.

|  | *Never Fallers*<br>*(n = 371)* | *Rare Fallers*<br>*(n = 178)* | *Frequent Fallers*<br>*(n = 56)* | *p(Never*<br>*≠Rare)* | *p(Rare*<br>*≠Frequent)* |
| --- | --- | --- | --- | --- | --- |
| *Sex: Male* | 261 (70.4%) * | 97 (55.5%) | 43 (76.8%) * | <0.001 | 0.01 |
| *Age (y)* | 62.5 (9.4) * | 62.9 (9.8) | 63.5 (8.7) | 0.003 | 0.028 |
| *Years since diagnosis* | 3.1 (3.0) * | 6.5 (4.5) | 10.1 (3.6) * | <m.p. | <0.001 |
| *Race: White* | 344 (93.5%) | 167 (95.4%) | 54 (96.4%) | 0.71 | 0.87 |
| *Ethnicity: Not Hispanic or Latino* | 354 (96.6%) * | 166 (92.3%) | 55 (98.2%) | 0.005 | 0.45 |
| *Education years* | 16 (2.8) | 16 (2.9) | 16.1 (2.4) | 0.51 | 0.82 |
| *BMI* | 27.2 (25.3) | 25.3 (4.1) | 26.6 (4.4) | 0.23 | 0.47 |
| *LEDD (mg)* | 294 (384) | 587 (572) | 829 (524) | 0.083 | 0.54 |
| *S&E* | 92.5 (7) | 86.8 (11.5) | 70.3 (17.3) * | 0.032 | <m.p. |
| *NSD-ISS* | 321 | 153 | 49 |  |  |
| *≤ Stage 3* | 264 (82.2%) * | 90 (58.8%) | 10 (20.4%) * | 0.002 | 0.002 |
| *> Stage 3* | 57 (17.8%) * | 63 (41.2%) | 39 (79.6%) * | 0.002 | 0.002 |
| *2a* | 1 (0.3%) | 0 | 0 |  |  |
| *2b* | 57 (17.8%) | 13 (8.5%) | 0 |  |  |
| *3* | 206 (64.2%) | 77 (50.3%) | 10 (20.4%) |  |  |
| *4* | 54 (16.8%) | 58 (37.9%) | 27 (55.1%) |  |  |
| *5* | 3 (0.9%) | 5 (3.3%) | 11 (22.4%) |  |  |
| *6* | 0 | 0 | 1 (2.0%) |  |  |
| *Not NSD-ISS* | 22 | 13 | 7 |  |  |

BMI, Body Mass Index; ED, Emergency Department; LEDD, Levodopa Equivalent Daily Dose; NSD-ISS, Neuronal Synuclein Disease-Integrated Staging System; S&E, Modified Schwab and England Activities of Daily Living Scale. Values are *n* or mean (with corresponding % or SD), as appropriate, \*: statistically significant from rare, $p < 0.016$ (BH threshold for FDR 0.05). m.p. = machine precision.

#### Motor and non-motor differences between never, rare, and frequent fallers

Frequent fallers exhibited significantly worse motor and non-motor profiles compared to rare and never fallers (Table 2). Motor severity, including higher H&Y stages and PIGD scores in both ON and OFF states, was greatest in frequent fallers (eFigure 2). Total MDS-UPDRS scores and Part II and III sub scores were also elevated in this group, while never fallers showed milder impairment. Differences between never and rare fallers were limited, reaching significance primarily for PIGD scores. MDS-UPDRS-Part IV scores were higher in frequent fallers compared to rare fallers, but did not differ significantly between never and rare fallers.

**Table 2:**
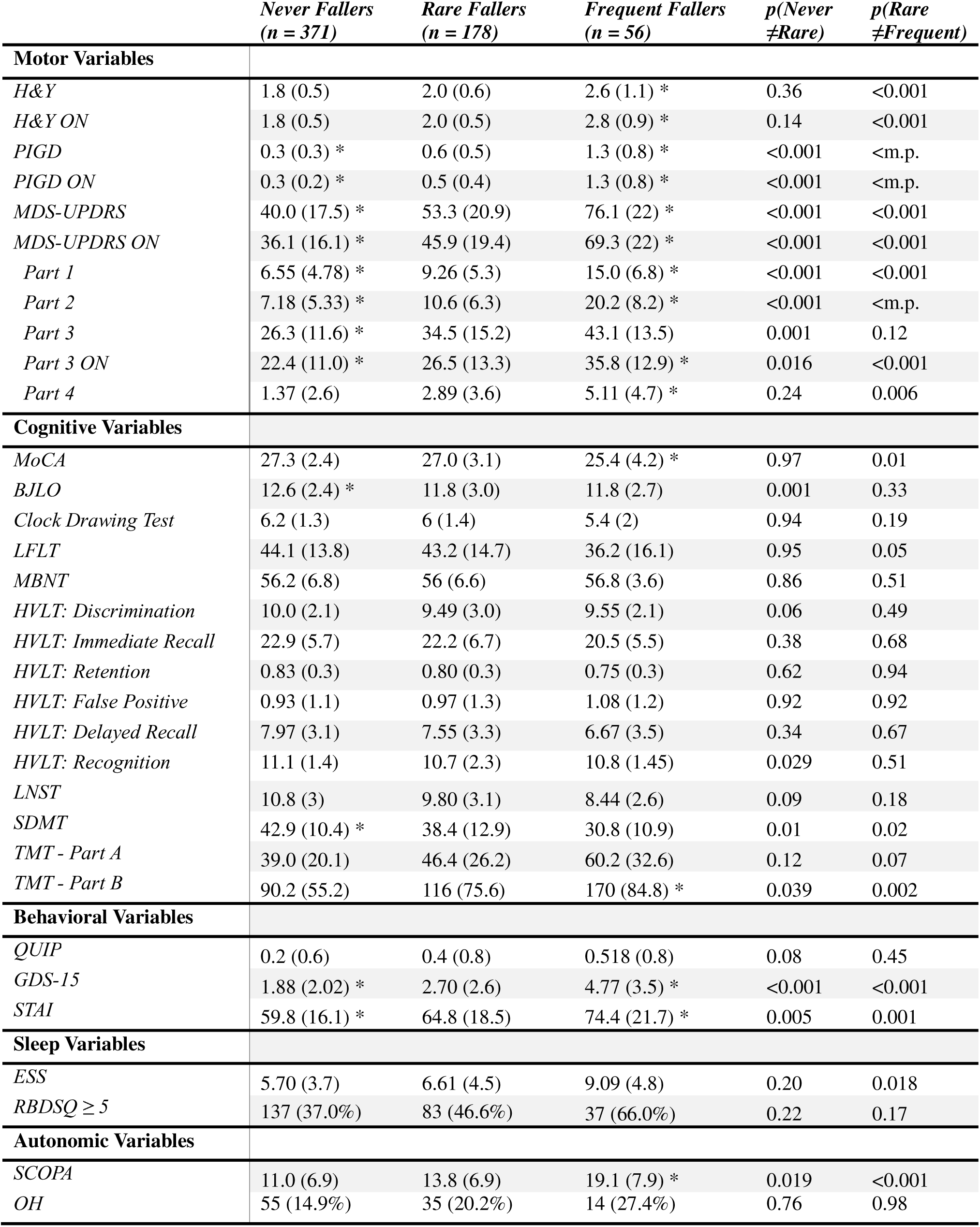
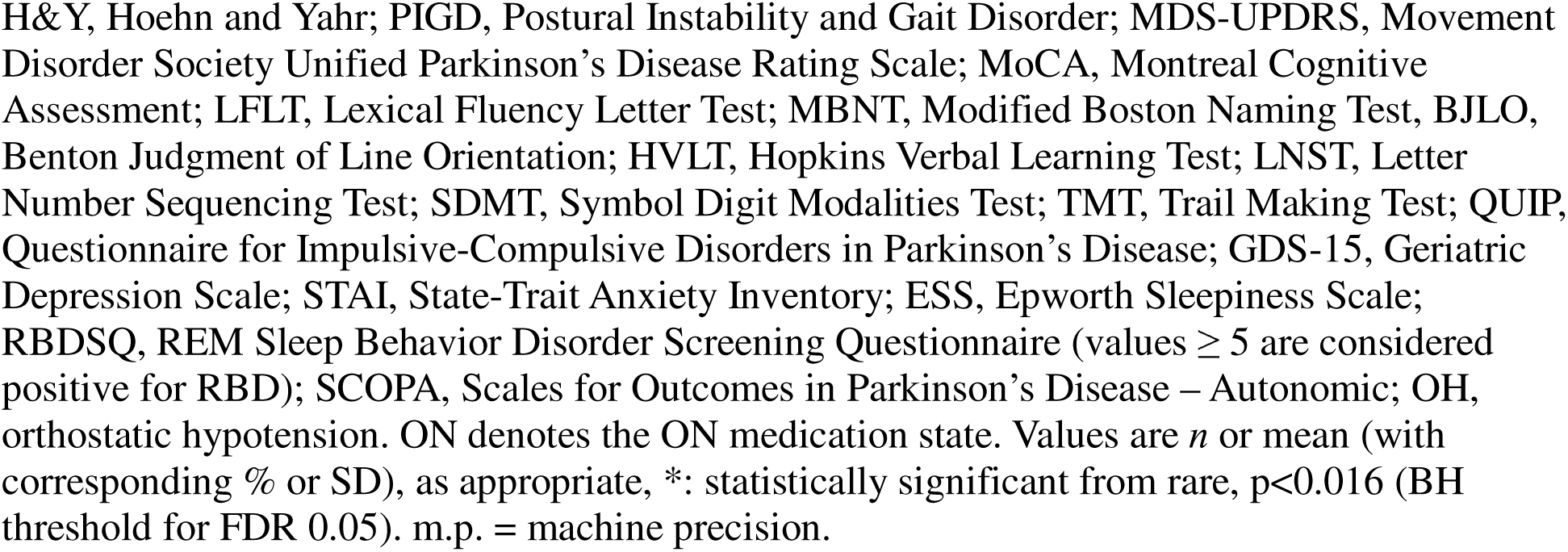
Motor and Non-motor Differences Between Never, Rare, and Frequent Fallers in the Cross-Sectional PD Cohort.

|  | *Never Fallers*<br>*(n = 371)* | *Rare Fallers*<br>*(n = 178)* | *Frequent Fallers*<br>*(n = 56)* | *p(Never*<br>*≠Rare)* | *p(Rare*<br>*≠Frequent)* |
| --- | --- | --- | --- | --- | --- |
| **Motor Variables** |  |  |  |  |  |
| *H&Y* | 1.8 (0.5) | 2.0 (0.6) | 2.6 (1.1) * | 0.36 | <0.001 |
| *H&Y ON* | 1.8 (0.5) | 2.0 (0.5) | 2.8 (0.9) * | 0.14 | <0.001 |
| *PIGD* | 0.3 (0.3) * | 0.6 (0.5) | 1.3 (0.8) * | <0.001 | <m.p. |
| *PIGD ON* | 0.3 (0.2) * | 0.5 (0.4) | 1.3 (0.8) * | <0.001 | <m.p. |
| *MDS-UPDRS* | 40.0 (17.5) * | 53.3 (20.9) | 76.1 (22) * | <0.001 | <0.001 |
| *MDS-UPDRS ON* | 36.1 (16.1) * | 45.9 (19.4) | 69.3 (22) * | <0.001 | <0.001 |
| *Part 1* | 6.55 (4.78) * | 9.26 (5.3) | 15.0 (6.8) * | <0.001 | <0.001 |
| *Part 2* | 7.18 (5.33) * | 10.6 (6.3) | 20.2 (8.2) * | <0.001 | <m.p. |
| *Part 3* | 26.3 (11.6) * | 34.5 (15.2) | 43.1 (13.5) | 0.001 | 0.12 |
| *Part 3 ON* | 22.4 (11.0) * | 26.5 (13.3) | 35.8 (12.9) * | 0.016 | <0.001 |
| *Part 4* | 1.37 (2.6) | 2.89 (3.6) | 5.11 (4.7) * | 0.24 | 0.006 |
| **Cognitive Variables** |  |  |  |  |  |
| *MoCA* | 27.3 (2.4) | 27.0 (3.1) | 25.4 (4.2) * | 0.97 | 0.01 |
| *BJLO* | 12.6 (2.4) * | 11.8 (3.0) | 11.8 (2.7) | 0.001 | 0.33 |
| *Clock Drawing Test* | 6.2 (1.3) | 6 (1.4) | 5.4 (2) | 0.94 | 0.19 |
| *LFLT* | 44.1 (13.8) | 43.2 (14.7) | 36.2 (16.1) | 0.95 | 0.05 |
| *MBNT* | 56.2 (6.8) | 56 (6.6) | 56.8 (3.6) | 0.86 | 0.51 |
| *HVLT: Discrimination* | 10.0 (2.1) | 9.49 (3.0) | 9.55 (2.1) | 0.06 | 0.49 |
| *HVLT: Immediate Recall* | 22.9 (5.7) | 22.2 (6.7) | 20.5 (5.5) | 0.38 | 0.68 |
| *HVLT: Retention* | 0.83 (0.3) | 0.80 (0.3) | 0.75 (0.3) | 0.62 | 0.94 |
| *HVLT: False Positive* | 0.93 (1.1) | 0.97 (1.3) | 1.08 (1.2) | 0.92 | 0.92 |
| *HVLT: Delayed Recall* | 7.97 (3.1) | 7.55 (3.3) | 6.67 (3.5) | 0.34 | 0.67 |
| *HVLT: Recognition* | 11.1 (1.4) | 10.7 (2.3) | 10.8 (1.45) | 0.029 | 0.51 |
| *LNST* | 10.8 (3) | 9.80 (3.1) | 8.44 (2.6) | 0.09 | 0.18 |
| *SDMT* | 42.9 (10.4) * | 38.4 (12.9) | 30.8 (10.9) | 0.01 | 0.02 |
| *TMT - Part A* | 39.0 (20.1) | 46.4 (26.2) | 60.2 (32.6) | 0.12 | 0.07 |
| *TMT - Part B* | 90.2 (55.2) | 116 (75.6) | 170 (84.8) * | 0.039 | 0.002 |
| **Behavioral Variables** |  |  |  |  |  |
| *QUIP* | 0.2 (0.6) | 0.4 (0.8) | 0.518 (0.8) | 0.08 | 0.45 |
| *GDS-15* | 1.88 (2.02) * | 2.70 (2.6) | 4.77 (3.5) * | <0.001 | <0.001 |
| *STAI* | 59.8 (16.1) * | 64.8 (18.5) | 74.4 (21.7) * | 0.005 | 0.001 |
| **Sleep Variables** |  |  |  |  |  |
| *ESS* | 5.70 (3.7) | 6.61 (4.5) | 9.09 (4.8) | 0.20 | 0.018 |
| *RBDSQ ≥ 5* | 137 (37.0%) | 83 (46.6%) | 37 (66.0%) | 0.22 | 0.17 |
| **Autonomic Variables** |  |  |  |  |  |
| *SCOPA* | 11.0 (6.9) | 13.8 (6.9) | 19.1 (7.9) * | 0.019 | <0.001 |
| *OH* | 55 (14.9%) | 35 (20.2%) | 14 (27.4%) | 0.76 | 0.98 |

H&Y, Hoehn and Yahr; PIGD, Postural Instability and Gait Disorder; MDS-UPDRS, Movement Disorder Society Unified Parkinson's Disease Rating Scale; MoCA, Montreal Cognitive Assessment; LFLT, Lexical Fluency Letter Test; MBNT, Modified Boston Naming Test, BJLO, Benton Judgment of Line Orientation; HVL, Hopkins Verbal Learning Test; LNST, Letter Number Sequencing Test; SDMT, Symbol Digit Modalities Test; TMT, Trail Making Test; QUIP, Questionnaire for Impulsive-Compulsive Disorders in Parkinson's Disease; GDS-15, Geriatric Depression Scale; STAI, State-Trait Anxiety Inventory; ESS, Epworth Sleepiness Scale; RBDSQ, REM Sleep Behavior Disorder Screening Questionnaire (values $\geq 5$ are considered positive for RBD); SCOPA, Scales for Outcomes in Parkinson's Disease – Autonomic; OH, orthostatic hypotension. ON denotes the ON medication state. Values are *n* or mean (with corresponding % or SD), as appropriate, \*: statistically significant from rare, $p < 0.016$ (BH threshold for FDR 0.05). m.p. = machine precision.

Non-motor symptoms followed a similar gradient (eFigure 3). Frequent fallers had higher MDS-UPDRS Part I scores, greater depressive and anxiety symptoms, and worse autonomic dysfunction, particularly in gastrointestinal, cardiovascular, urinary, and thermoregulatory domains. Cognitively, frequent fallers performed worse on MoCA, processing speed (e.g., SDMT), visuospatial judgment (e.g., BJLO), and executive function (e.g., TMT-B), while other domains showed no group differences. Although OH was more common in frequent fallers, this association was not statistically significant after adjusting for confounders.

Female PwP had milder PD symptoms, scoring five points lower on the total MDS-UPDRS ON exam, and specifically 1.5 points lower on MDS-UPDRS Part II, than their male counterparts (eTable 6). Notably, MDS-UPDRS Part III, PIGD, and H&Y were not significantly different between sexes on either ON or OFF states. Cognitively, female PwP outperformed males on several domains, including verbal learning and memory (e.g., HVLT), verbal fluency (e.g., LFLT), processing speed (e.g., SDMT), and cognitive flexibility and visual attention (e.g., TMT Parts A and B). In contrast, male PwP performed better on visuospatial judgment (e.g., BJLO). Finally, female PwP were less likely to have sleep disturbances, with a lower percentage of RBDSQ ≥ 5 rates and a lower average ESS.

#### Fall-related outcome differences between rare and frequent fallers

Frequent fallers were significantly more likely than rare fallers to report any fall-related injury (66.1% versus 39.9%), including head injuries (33.9% versus 11.8%), lacerations (25% versus 6.7%), and multiple injuries (33.9% versus 8.4%; Table 3). They also reported higher rates of emergency department visits (50% versus 20.2%) due to fall-related injuries (eFigure 4). Female PwP were more likely to report any fracture (10.3% versus 4.24%), particularly upper extremity fractures (5.4% versus 1.8%) than their male counterparts (eFigure 5). Differences in hip fractures between female and male PwP (3.4% versus 0.8%) did not reach statistical significance (eTable 6).

**Table 3:** Outcome Differences Between Rare and Frequent Fallers in the Cross-Sectional PD Cohort.

|  | *Rare Fallers (n = 178)* | *Frequent Fallers (n = 56)* | *p(Rare ≠Frequent)* |
| --- | --- | --- | --- |
| **Injury** |  |  |  |
| *Any Injury* | 71 (39.9%) | 37 (66.1%) | 0.012* |
| *Any Fracture* | 24 (13.5%) | 14 (25.0%) | 0.25 |
| *Hip Fracture* | 7 (3.9%) | 3 (5.4%) | 0.82 |
| *Upper Extremity Fracture* | 10 (5.6%) | 8 (14.3%) | 0.075 |
| *Skull Fracture* | 2 (1.1%) | 0 (0%) | 1.0 |
| *Other Fracture* | 11 (6.2%) | 7 (12.5%) | 0.36 |
| *Head Injury* | 21 (11.8%) | 19 (33.9%) | 0.016 * |
| *Laceration* | 12 (6.7%) | 14 (25.0%) | 0.019 |
| *Other Injury* | 34 (19.1%) | 21 (37.5%) | 0.005 * |
| *Multiple Injuries* | 15 (8.4%) | 19 (33.9%) | <0.001 * |
| **Healthcare Utilization** |  |  |  |
| *Any Healthcare Use* | 55 (30.9%) | 34 (60.7%) | 0.023 |
| *Outpatient Visit* | 32 (18.0%) | 24 (42.9%) | 0.033 |
| *ED Visit* | 36 (20.2%) | 28 (50.0%) | 0.005 * |
| *Hospitalization* | 11 (6.2%) | 9 (16.1%) | 0.33 |
| *Surgery* | 6 (3.4%) | 7 (12.5%) | 0.095 |
| *Institutionalization* | 2 (1.1%) | 0 (0%) | 1.0 |

BMI, Body Mass Index; ED, Emergency Department; LEDD, Levodopa Equivalent Daily Dose; S&E, Modified Schwab and England Activities of Daily Living Scale. Values are *n* or mean (with corresponding % or SD), as appropriate, \*: statistically significant from rare, $p < 0.016$ (BH threshold for FDR 0.05). m.p. = machine precision.

## DISCUSSION

In this large, multicohort study, we found that falls were significantly more common in PwP, with odds about 1.5-fold higher than PAS and over fourfold higher than HC. Fall-related injuries and healthcare utilization were also more frequent, largely explained by increased fall frequency rather than PD diagnosis alone, and increased in parallel with years since diagnosis and NSD-ISS stage. Frequent fallers had more severe motor and non-motor symptoms compared to rare or never fallers. Notably, female sex emerged as an independent predictor of falls and related morbidity, despite a milder overall clinical phenotype. Overall, our study provides estimates of fall frequency across a wide range of years since diagnosis, identifying associated clinical and biological features, and demonstrating the utility of NSD-ISS staging in stratifying fall-related vulnerability in PD

Falls occurred in over one-fourth of visits among PwP and increased with both years since diagnosis and advancing disability, as indicated by NSD-ISS stage 4 or higher. By 7 years post-diagnosis, one-third of visits reported a fall, and about one-tenth involved frequent falls. A similar trend was observed for fall-related injuries and healthcare utilization, suggesting that longer disease duration contributes not only to fall risk but also to their severity and consequences. Notably, approximately 15% of PwP had already experienced a fall at the time of diagnosis. Although the injury rate at diagnosis was relatively low, this finding supports the need for fall screening from the time of diagnosis, as recommended in the American Academy of Neurology’s universal neurology quality measurement set.^22^ Early screening can inform both primary prevention strategies, such as physical therapy and home safety evaluations, and secondary prevention efforts, including the use of assistive devices and bone health management.

Our findings should be interpreted in the context of the existing literature on falls in PD. In 22 prior studies, an average of 60.5% of individuals with PD reported at least one fall.^23^ Among community-dwelling individuals without dementia, similar to our study population, yearly fall rates have ranged from 21% to 65% when followed for up to 12 months.^24–31^ Longitudinal studies have also described high rates of frequent falls. For example, in the Norwegian ParkWest study,^32^ 46% of participants with newly diagnosed PD experienced frequent falls by a median follow-up time of six years. These rates are higher than those observed in our study, which may reflect length-of-time bias. Individuals with slower disease progression are more likely to remain in long-term follow-up, and thus overrepresented in our sample, potentially leading to an underestimation of the true frequency of falls. The relationship between fall frequency and NSD-ISS stage further supports the validity of this recently proposed staging system. Although falls are not incorporated into the NSD-ISS criteria, fall frequency increased consistently across higher stages. This aligns the transition from NSD-ISS stage 3 to 4 as an inflexion point linked to a meaningful, patient-centered outcome and reinforces the framework of increasing levels of disability at increasing stages.^33^

We also observed that frequent fallers had longer years since diagnosis, higher NSD-ISS stages, greater functional impairment, and more severe motor and non-motor symptoms compared to rare or never fallers. They showed higher H&Y stages, PIGD scores, and MDS-UPDRS total and subscale scores, including greater motor complications on Part IV. In contrast, LEDD did not differ significantly across groups, suggesting dopaminergic dose alone may not explain fall risk.

Instead, treatment-related complications, rather than absolute dose, may contribute to greater fall vulnerability in frequent fallers. Cognitive decrements were most evident in processing speed, executive function, and visuospatial perception, with other domains showing no differences, underscoring the selective nature of cognitive vulnerability in PwP.^11^ OH rates trended higher with fall frequency but were confounded by age and disease duration, whereas SCOPA cardiovascular scores were significantly worse in frequent fallers, indicating patient-reported OH symptoms may better capture fall risk than objective measures. This mismatch between objective and subjective OH symptoms and fall risk may be explained by differences in immediate versus delayed or symptomatic versus asymptomatic OH,^34^ which may contribute differently to fall risk.

Sex was a consistent predictor of fall risk and outcomes. Female PwP, despite lower LEDD needs, milder motor symptoms, and better cognitive performance, were significantly more likely than males to report falls, injuries, and healthcare use, even after adjustment for age, disease duration, and fall frequency. This excess risk was most evident for rare falls and upper extremity fractures; hip fractures did not reach significance due to low counts. Ongoing trials of osteoporosis treatment, a condition disproportionately affecting women, may help mitigate fracture risk in PD.^35^ Several factors may underlie sex differences. Biomechanically, females’ lower center of mass may enhance static stability but impair recovery from imbalance, while males’ higher center of mass may predispose to imbalance yet favor corrective movements.^36^ Female PwP also show higher fall risk independent of age,^37,38^ despite fewer axial symptoms than males,^39^ and may be additionally influenced by urinary incontinence,^40^ traditional gendered expectations (e.g., domestic tasks such as cooking and cleaning),^41^ a lower socioeconomic status,^42^ or a greater willingness to report falls and seek healthcare than their male counterparts.^43^ These findings highlight a disconnect between conventional severity markers and real-world vulnerability in women with PD, underscoring the need for sex-specific interventions.

Our study has several limitations. Differential withdrawal of participants with greater disability, including falls, may have led to underestimates in advanced PD. We did not evaluate the effect of socioeconomic status, comorbidities, or polypharmacy, all recognized fall risk factors. Fall definitions relied on self-report from annual questionnaires rather than fall diaries (i.e., the gold standard), raising the possibility of underreporting due to recall or social desirability bias (i.e., underreporting to appear healthier). To reduce misclassification, we required consecutive years of fall reporting in our cross-sectional analysis. Although our data span over 14 years since diagnosis, the mean duration for unique participants was 5 years, reflecting the later introduction of the fall questionnaire. Earlier enrollees generally had longer disease duration, while later enrollees had shorter, which we addressed by adjusting for years since diagnosis.

## CONCLUSION

Our findings highlight the high burden of falls and fall-related outcomes in PD, driven by disease progression and amplified by sex-specific vulnerabilities. Fall risk reflected motor, cognitive, behavioral, and autonomic dysfunction, supporting a multifactorial model of susceptibility. The strong link between fall frequency and NSD-ISS stage further validates this staging system as a clinical tool. These results highlight the importance of early screening, tailored prevention strategies, and attention to sex-specific risks. Fall frequency may serve as a practical marker of disease progression and a target for interventions to reduce morbidity in PD.

## Financial Disclosure/Conflict of Interest

none

## Funding

This work was supported by the National Institute of Health (1T32NS091006-10 and 5T32NS091006-10), the Institute for Translational Medicine and Therapeutics of the Perelman School of Medicine at the University of Pennsylvania, and the Michael J. Fox Foundation (Write Now! Award). Study sponsors had no role in the data for the present study.

## Supporting information

eTable

## Data Availability

Data, protocol, and materials may be requested from the PPMI at https://www.ppmi-info.org.

## Acknowledgements

Protocol information for The Parkinson’s Progression Markers Initiative (PPMI) Clinical - Establishing a Deeply Phenotyped PD Cohort AM 3.2. can be found on protocols.io or by following this link: ttps://dx.doi.org/10.17504/protocols.io.n92ldmw6ol5b/v2. PPMI – a public-private partnership – is funded by the Michael J. Fox Foundation for Parkinson’s Research and funding partners, including 4D Pharma, Abbvie, AcureX, Allergan, Amathus Therapeutics, Aligning Science Across Parkinson’s, AskBio, Avid Radiopharmaceuticals, BIAL, BioArctic, Biogen, Biohaven, BioLegend, BlueRock Therapeutics, Bristol-Myers Squibb, Calico Labs, Capsida Biotherapeutics, Celgene, Cerevel Therapeutics, Coave Therapeutics, DaCapo Brainscience, Denali, Edmond J. Safra Foundation, Eli Lilly, Gain Therapeutics, GE HealthCare, Genentech, GSK, Golub Capital, Handl Therapeutics, Insitro, Jazz Pharmaceuticals, Johnson & Johnson Innovative Medicine, Lundbeck, Merck, Meso Scale Discovery, Mission Therapeutics, Neurocrine Biosciences, Neuron23, Neuropore, Pfizer, Piramal, Prevail Therapeutics, Roche, Sanofi, Servier, Sun Pharma Advanced Research Company, Takeda, Teva, UCB, Vanqua Bio, Verily, Voyager Therapeutics, the Weston Family Foundation and Yumanity Therapeutics.

## Code Availability

Code is available at https://github.com/KHefter/PPMIFallsAndInjury.

## Data Availability

Data used in the preparation of this article were obtained on 2024-12-11 and 2025-01-10 from the Parkinson’s Progression Markers Initiative (PPMI) database (https://www.ppmi-info.org/access-data-specimens/download-data), RRID:SCR_006431. For up-to-date information on the study, visit http://www.ppmi-info.org. This analysis used data openly available from the PPMI (Tier 1 data).

## Authors’ disclosures

JAV: is an employee of the University of Pennsylvania and received funding support from the National Institutes of Health (1T32NS091006 10), the Institute for Translational Medicine and Therapeutics of the Perelman School of Medicine at the University of Pennsylvania, and the Michael J. Fox Foundation; honoraria from the International Parkinson and Movement Disorder Society; and consultancy fees from the University City Science Center.

KH: is a graduate student at the University of Pennsylvania and received funding from the National Institutes of Health (5T32NS091006-10).

DEL: is an employee of the University of Iowa.

MTD: is an employee of the University of Pennsylvania and received funding support from the National Institutes of Health (F30-AG074524).

AE: is an employee of the University of Pennsylvania and received funding support from the National Institutes of Health and The Patient-Centered Outcomes Research Institute.

BL: is an employee of the University of Pennsylvania and received funding support from NINDS (DP1-NS-122038-01), The Small Lake Foundation, Jonathan and Bonnie Rothberg, Neil and Barbara Smit.

DSB: is a faculty member at the University of Pennsylvania.

AS: is a faculty member at the University of Pennsylvania; consultancy fees in the past year from Acadia, Eli Lilly and Co, Neurocrine, Theravance, Cerevance, Spark/Roche, Boerhinger-Ingelheim, Wave Life Sciences, Inhibikase, Prevail, Mitzubishi and Alertity Therapeutics. He has served on DSMBs for the Huntington Study Group and The Healey ALS Consortium (Massachusetts General Hospital). He has received grant funding from the Michael J. Fox Foundation, NIA and NINDS.

## Authors’ roles

JAV: Design, execution, analysis, writing, and editing of the final version of the manuscript. KH: Design, execution, analysis, writing, and editing of the final version of the manuscript. DEL: Editing of the final version of the manuscript.

MTD: Editing of the final version of the manuscript

AE: Analysis and editing of the final version of the manuscript. BL: Editing of the final version of the manuscript

DSB: Analysis and editing of the final version of the manuscript

AS: Design, execution, analysis, and editing of the final version of the manuscript.

## Notes

### Competing Interest Statement

The authors have declared no competing interest.

### Author Declarations

The study was approved by the internal review boards of all participating PPMI clinical sites.

### Summary of Updates

Updated code to reflect adjustment by age at time of visit and not baseline. Updated manuscript to reflect changes.

