## Supplementary material for "Fall Frequency, Risk Factors, and Outcomes in Parkinson’s Disease: A Cross-Sectional Analysis": eTable

**Contents:**

**eTable 1: Fall Frequency Classification Based on Unified Parkinson’s Disease Rating Scale Items 13 and 14**

**eTable 2: Frequency of Falls, Injury, and Healthcare Utilization Between PD, PAS, and HC**

**eTable 3: Effect of Cohort Between PD and PAS on Injury and Healthcare Utilization**

**eTable 4: Effect of Fall Frequency and Sex on Injury and Healthcare Utilization Measures in Fallers with PD and PAS**

**eTable 5: Falls, Injury, and Healthcare Utilization across NSD-ISS Stages in PD Patients**

**eTable 6: Sex-Based Differences in the Cross-sectional PD Cohort**

**eFigure 1: Sampling without Replacement for Cross-sectional Analysis**

**eFigure 2: Motor Differences between Never, Rare, And Frequent Fallers in PD**

**eFigure 3: Non-Motor Differences Between Never, Rare, and Frequent Fallers in PD**

**eFigure 4: Outcome Differences between Rare and Frequent Fallers in PD**

**eFigure 5: Sex-based Differences in the Cross-Sectional PD Cohort**

**eAppendix: PPMI Author List**

| *Fall frequency* | *Item 13 score* | *Combination* | *Item 14 score* |
| --- | --- | --- | --- |
| *None* | 0 = none | AND | ≤2 (0 = none; 1 = rare freezing when walking; 2 = occasional freezing when walking) |
| *Rare* | 1 = rare falling | AND | ≤2 (0 = none; 1 = rare freezing when walking; 2 = occasional freezing when walking) |
| *Frequent* | ≥2 (2 = Occasionally falls; 3 = falls an average of once daily; 4 = falls more than once daily) | OR | ≥3 (3 = occasionally falls from freezing; 4 = frequent falls from freezing) |

**eTable 1: Fall Frequency Classification Based on Unified Parkinson’s Disease Rating Scale Items 13 and 14**

Fall frequency was determined by the following response combinations on Unified Parkinson’s Disease Rating Scale items 13 and 14.

**eTable 2: Frequency of Falls, Injury, and Healthcare Utilization Between PD, PAS, and HC**

|  | **PD**  **(n = 2374 visits)** | **PAS**  **(n = 3848 visits)** | | | **HC**  **(n = 757 visits)** | | |
| --- | --- | --- | --- | --- | --- | --- | --- |
|  | **N (%)** | **N (%)** | **OR**  **(95% CI)** | **p** | **N (%)** | **OR**  **(95% CI)** | **p** |
| **Falls** | | | | | | | |
| Any Falls | 677 (28.5%) | 850 (22.1%) | 1.66 *  (1.46, 1.87) | <0.001 | 80 (10.6%) | 4.03 *  (3.14, 5.23) | <0.001 |
| Rare Falls | 514 (21.7%) | 806 (20.9%) | 1.22 *  (1.06, 1.38) | 0.003 | 78 (10.3%) | 2.75 *  (2.14, 3.59) | <0.001 |
| Frequent Falls | 163 (6.9%) | 44 (1.1%) | 6.49 *  (4.66, 9.24) | <0.001 | 2 (0.3%) | 33.2 *  (10.5, 201.1) | <0.001 |
| **Injuries** | | | | | | | |
| Any Injury | 264 (11.1%) | 303 (7.9%) | 1.70 *  (1.42, 2.04) | <0.001 | 33 (4.4%) | 3.26 *  (2.27, 4.85) | <0.001 |
| Multiple Injuries | 70 (2.9%) | 65 (1.7%) | 1.85 *  (1.31, 2.64) | <0.001 | 5 (0.7%) | 6.32 *  (2.27, 18.2) | 0.001 |
| Any Fracture | 80 (3.4%) | 96 (2.5%) | 1.55 *  (1.14, 2.11) | 0.005 | 10 (1.3%) | 3.36 *  (1.80, 7.01) | 0.005 |
| Hip Fracture | 18 (0.8%) | 32 (0.8%) |  | NS | 2 (0.3%) |  | NS |
| Upper Extremity Fracture | 32 (1.3%) | 31 (0.8%) | 2.03 *  (1.23, 3.38) | 0.006 | 6 (0.8%) | 2.14  (0.94, 5.74) | 0.09 |
| Skull Fracture | 4 (0.2%) | 2 (0.1%) |  | NS | 0 (0%) |  | NS |
| Other Fracture | 35 (1.5%) | 37 (1.0%) | 1.61  (1.00, 2.56) | 0.049 | 2 (0.3%) | 7.00 *  (2.11, 43.3) | 0.008 |
| Head Injury | 85 (3.6%) | 82 (2.1%) | 1.78 *  (1.29, 2.42) | <0.001 | 8 (1.1%) | 4.60 *  (2.33, 10.5) | <0.001 |
| Lacerations | 48 (2.0%) | 20 (0.5%) | 3.92 *  (2.34, 6.82) | <0.001 | 5 (0.7%) | 4.03 *  (1.73, 11.7) | 0.004 |
| Other Injury | 139 (5.9%) | 177 (4.6%) | 1.50 *  (1.19, 1.89) | <0.001 | 18 (2.4%) | 2.85 *  (1.77, 4.86) | <0.001 |
| **Healthcare Utilization** | | | | | | | |
| Any HC Use | 202 (8.5%) | 226 (5.9%) | 1.71 *  (1.39, 2.09) | <0.001 | 23 (3.0%) | 3.81 *  (2.48, 6.12) | <0.001 |
| Outpatient Visit | 112 (4.7%) | 149 (3.9%) | 1.39 *  (1.07, 1.79) | 0.012 | 13 (1.7%) | 3.61 *  (2.08, 6.82) | <0.001 |
| ED Visit | 138 (5.8%) | 130 (3.4%) | 1.99 *  (1.54, 2.56) | <0.001 | 15 (2.0%) | 3.97 *  (2.37, 7.16) | <0.001 |
| Hospitalization | 40 (1.7%) | 27 (0.7%) | 2.46 *  (1.50, 4.10) | <0.001 | 7 (0.9%) | 2.54  (1.19, 6.33) | 0.026 |
| Surgery | 29 (1.2%) | 34 (0.9%) |  | NS | 8 (1.1%) |  | NS |
| Institutionalization | 5 (0.2%) | 0 (0%) |  | NS | 1 (0.1%) |  | NS |

ED, Emergency Department; HC, Healthy Control; NS, not significant on the Wald test; OR, Odds ratio; PD, Parkinson’s disease; PAS, Prodromal alpha-synucleinopathy. *: statistically significant from PD, p<0.028 (BH threshold for FDR 0.05). Model also conditioned on age and sex (odds ratios/significance not shown). OR calculated as PD/another cohort. OR only calculated for variables with models for which cohort membership provided a significant improvement (p<0.05 using Wald test) over models without cohort membership.

**eTable 3: Effect of Cohort Between PD and PAS on Injury and Healthcare Utilization**

|  | **PD**  **(n = 514 rare fall visits**  **n = 163 frequent fall visits)** | **PAS**  **(n = 806 rare fall visits**  **n = 44 frequent fall visits)** |  | |
| --- | --- | --- | --- | --- |
|  | ***N (%)*** | ***N (%)*** | ***OR (95% CI)*** | ***p*** |
| Any Injury | 264 (39.0%) | 303 (35.6%) | 1.16 (0.89, 1.40) | 0.35 |
| Fracture: | 80 (11.8%) | 96 (11.3%) | 1.01 (0.71, 1.42) | 0.97 |
| Hip Fracture | 18 (2.7%) | 32 (3.8%) | 0.75 (0.39, 1.39) | 0.36 |
| Upper Extremity Fracture | 32 (4.7%) | 31 (3.6%) | 1.26 (0.72, 2.18) | 0.41 |
| Skull Fracture | 4 (0.6%) | 2 (0.2%) | 2.50 (0.44, 19.14) | 0.31 |
| Other Fracture | 35 (5.2%) | 37 (4.4%) | 1.08 (0.65, 1.80) | 0.76 |
| Head Injury | 85 (12.6%) | 82 (9.6%) | 1.14 (0.80, 1.62) | 0.48 |
| Laceration | 48 (7.1%) | 20 (2.4%) | 2.17 * (1.23, 3.94) | 0.008 |
| Other Injury | 139 (20.5%) | 177 (20.8%) | 0.96 (0.73, 1.25) | 0.76 |
| Multiple Injuries | 70 (10.3%) | 65 (7.64%) | 1.07 (0.72, 1.58) | 0.75 |
| Any HC Use | 202 (29.8%) | 226 (26.6%) | 1.07 (0.83, 1.36) | 0.61 |
| Doctor Visit | 112 (16.5%) | 149 (17.5%) | 0.83 (0.62, 1.11) | 0.21 |
| ED Visit | 138 (20.4%) | 130 (15.3%) | 1.22 (0.92, 1.65) | 0.17 |
| Hospitalization | 40 (5.9%) | 27 (3.2%) | 1.59 (0.92, 2.75) | 0.09 |
| Surgery | 29 (4.3%) | 34 (4.0%) | 0.96 (0.54, 1.66) | 0.88 |
| Institutionalization | 5 (0.7%) | 0 (0%) | n/a | n/a |

Differences in injury and healthcare utilization measures for fallers with PD and PAS. OR, odds ratio: PD/PAS. Parkinson’s disease; PAS, Prodromal alpha-synucleinopathy. ORs calculated from a model conditioned on age, fall frequency, sex, and cohort. Results for OR based on fall frequency and sex are in Supplementary Table 4. *: statistically significant difference between PD / PAS, p<0.024 (BH threshold for FDR 0.05). m.p. = machine precision.

**eTable 4: Effect of Fall Frequency and Sex on Injury and Healthcare Utilization Measures in Fallers with PD and PAS**

|  | **Fall Frequency** | | | | **Sex** | | | |
| --- | --- | --- | --- | --- | --- | --- | --- | --- |
|  | ***Rare Fallers***  ***N (%)*** | ***Frequent Fallers***  ***N (%)*** | ***OR***  ***(95% CI)*** | ***p*** | ***Male***  ***N (%)*** | ***Female N (%)*** | ***OR***  ***(95% CI)*** | ***p*** |
| Total visits | 1320 | 207 |  |  | 727 | 800 |  |  |
| **Injuries** | | | | | | | | |
| Any Injury | 461 (34.9%) | 106 (51.2%) | 1.95 *  (1.42, 2.67) | <0.001 | 248 (34.1%) | 319 (39.9%) | 1.50 *  (1.20, 1.88) | <0.001 |
| Multiple Injuries | 94 (7.1%) | 41 (19.8%) | 2.88 *  (1,84, 4.44) | <0.001 | 68 (9.4%) | 67 (8.4%) | 1.27  (0.86, 1.87) | 0.22 |
| Any Fracture | 140 (10.6%) | 36 (17.4%) | 1.74 *  (1.12, 2.66) | 0.014 | 74 (10.2%) | 102 (12.8%) | 1.62 *  (1.15, 2.29) | 0.006 |
| Hip Fracture | 42 (3.2%) | 8 (3.9%) | 1.42  (0.58, 3.09) | 0.40 | 16 (2.2%) | 34 (4.3%) | 2.30 *  (1.23, 4.49) | 0.01 |
| Upper Extremity Fracture | 45 (3.4%) | 18 (8.7%) | 2.77 *  (1.47, 5.08) | 0.001 | 21 (2.9%) | 42 (5.3%) | 2.67 *  (1.51, 4.85) | 0.001 |
| Skull Fracture | 5 (0.4%) | 1 (0.5%) | 0.70  (0.03, 4.95) | 0.76 | 3 (0.4%) | 3 (0.4%) | 1.94  (0.32, 11.90) | 0.46 |
| Other Fracture | 59 (4.5%) | 13 (6.3%) | 1.23  (0.61, 2.31) | 0.54 | 39 (5.4%) | 33 (4.1%) | 0.88  (0.53, 1.46) | 0.62 |
| Head Injury | 129 (9.8%) | 38 (18.4%) | 1.74 *  (1.12, 2.64) | 0.01 | 89 (12.2%) | 78 (9.8%) | 1.00  (0.71, 1.42) | 0.98 |
| Laceration | 41 (3.1%) | 27 (13.0%) | 3.07 *  (1.76, 5.30) | <0.001 | 43 (5.9%) | 25 (3.1%) | 0.85  (0.49, 1.46) | 0.56 |
| Other Injury | 258 (19.5%) | 58 (28.0%) | 1.77*  (1.24, 2.51) | 0.02 | 130 (17.9%) | 186 (23.3%) | 1.46 *  (1.12, 1.91) | 0.005 |
| **HC Utilization** | | | | |  |  |  |  |
| Any HC Utilization | 336 (25.5%) | 92 (44.4%) | 2.27 *  (1.64, 3.14) | <0.001 | 193 (26.5%) | 235 (29.4%) | 1.48 *  (1.16, 1.89) | 0.002 |
| Outpatient Visit | 205 (15.5%) | 56 (27.1%) | 2.14 *  (1.47, 3.08) | <0.001 | 115 (15.8%) | 146 (18.3%) | 1.41 *  (1.06, 1.89) | 0.02 |
| ED Visit | 199 (15.1%) | 69 (33.3%) | 2.55 *  (1.79, 3.62) | <0.001 | 124 (17.1%) | 144 (18.0%) | 1.49 *  (1.11, 1.99) | 0.007 |
| Hospitalization | 49 (3.7%) | 18 (8.7%) | 1.75  (0.94, 3.17) | 0.07 | 37 (5.1%) | 30 (3.8%) | 1.11  (0.65, 1.90) | 0.68 |
| Surgery | 47 (3.6%) | 16 (7.7%) | 2.34 *  (1.21, 4.35) | 0.009 | 26 (3.6%) | 37 (4.6%) | 1.63  (0.95, 2.86) | 0.08 |
| Institutionalization | 4 (0.3%) | 1 (0.5%) | 0.76  (0.04, 5.38) | 0.82 | 3 (0.4%) | 2 (0.3%) | 1.07  (0.14, 6.78) | 0.94 |

HC, Healthcare; ED, Emergency Department; OR, Odds Ratio; Parkinson’s disease; PAS, Prodromal alpha-synucleinopathy. *: statistically significant difference between rare/frequent fallers or male/female, p<0.024 (BH threshold for FDR 0.05). m.p. = machine precision. Model conditioned on age (odds significance not shown). OR: frequent/rare; female/male.

**eTable 5: Falls, Injury, and Healthcare Utilization across NSD-ISS stages in PD patients**

| **NSD Stage** | **Total Visits** | **Fall Occurrences** | **Rare Falls** | **Frequent Falls** | **Any Injury** | **Any Healthcare Use** |
| --- | --- | --- | --- | --- | --- | --- |
| **2a** | 4 | 0 (0%) | 0 (0%) | 0 (0%) | 0 (0%) | 0 (0%) |
| **2b** | 258 | 22 (8.5%) | 22 (8.5%) | 0 (0%) | 8 (3.1%) | 5 (1.9%) |
| **3** | 1120 | 229 (20.4%) | 205 (18.3%) | 24 (2.1%) | 80 (7.1%) | 60 (5.4%) |
| **4** | 561 | 253 (45.1%) | 181 (32.3%) | 72 (12.8%) | 101 (18.0%) | 83 (14.8%) |
| **5** | 83 | 63 (75.9%) | 24 (28.9%) | 39 (47.0%) | 37 (44.6%) | 29 (34.9%) |
| **6** | 9 | 8 (88.8%) | 1 (11.1%) | 7 (77.8%) | 7 (77.8%) | 6 (66.7%) |

Values are counts with corresponding percentages.

**eTable 6: Sex-Based Differences in the Cross-Sectional PD Cohort**

|  | **Male**  **(n = 401)** | **Female**  **(n = 204)** | **p(Male ≠Female)** |
| --- | --- | --- | --- |
| Fall Occurrence | 140 (34.9%) | 94 (46.1%) * | 0.002 |
| Rare Faller | 97 (24.2%) | 81 (39.7%) * | <0.001 |
| Frequent Faller | 43 (10.7%) | 13 (6.4%) | 0.14 |
| BMI | 27.35 (11.92) | 25.14 (4.69) | 0.023 |
| LEDD (mg) | 464.68 (545.22) | 360.65 (367.34) * | 0.001 |
| S&E | 88.28 (12.36) | 89.80 (10.25) | 0.14 |
| NSD-ISS | 343 | 180 |  |
| ≤ Stage 3 | 234 (68.2%) | 130 (72.2%) | 0.20 |
| > Stage 3 | 109 (31.8%) | 50 (27.8%) |  |
| Not NSD-ISS | 29 | 13 |  |
| **Injury Variables** |  |  |  |
| Any Injury | 62 (15.5%) | 46 (22.5%) | 0.13 |
| Any Fracture | 17 (4.24%) | 21 (10.3%) * | 0.004 |
| Hip Fracture | 3 (0.75%) | 7 (3.43%) | 0.021 |
| Upper Extremity Fracture | 7 (1.75%) | 11 (5.39%) * | 0.014 |
| Head Injury | 28 (6.98%) | 12 (5.88%) | 0.58 |
| Laceration | 20 (4.99%) | 6 (2.94%) | 0.30 |
| Other Injury | 28 (6.98%) | 27 (13.2%) | 0.03 |
| Multiple Injuries | 21 (5.24%) | 13 (6.37%) | 0.32 |
| **Motor Variables** |  |  |  |
| H&Y | 1.91 (0.59) | 1.92 (0.63) | 0.75 |
| H&Y ON | 1.91 (0.61) | 1.88 (0.61) | 0.76 |
| PIGD | 0.44 (0.50) | 0.42 (0.41) | 0.98 |
| PIGD ON | 0.42 (0.62) | 0.39 (0.38) | 0.88 |
| MDS-UPDRS | 46.80 (21.89) | 44.62 (19.64) | 0.13 |
| MDS-UPDRS ON | 42.66 (20.41) | 38.63 (17.98) * | 0.016 |
| Part 1 | 8.21 (5.72) | 7.98 (5.72) | 0.65 |
| Part 2 | 9.94 (7.35) | 8.31 (6.28) * | 0.003 |
| Part 3 | 30.00 (14.26) | 29.26 (12.80) | 0.36 |
| Part 3 ON | 25.30 (12.57) | 23.18 (11.84) | 0.05 |
| Part 4 | 2.29 (3.56) | 2.45 (3.36) | 0.70 |
| **Cognitive & Behavioral Variables** |  |  |  |
| BJLO | 12.51 (2.55) | 11.76 (2.64) * | 0.004 |
| LFLT | 42.07 (13.95) | 45.27 (15.07) * | 0.016 |
| QUIP | 0.33 (0.74) | 0.20 (0.56) | 0.019 |
| SDMT | 39.47 (12.04) | 42.83 (10.82) * | <0.001 |
| TMT - Part A | 44.91 (25.97) | 39.14 (19.18) * | 0.002 |
| TMT - Part B | 110.09 (71.95) | 93.42 (59.36) * | 0.001 |
| **Sleep Variables** |  |  |  |
| ESS | 6.78 (4.17) | 5.29 (3.90) * | <0.001 |
| RBDSQ ≥ 5 | 186 (46.4%) | 71 (35.0%) * | 0.007 |
| **Autonomic Variables** |  |  |  |
| SCOPA | 12.95 (7.47) | 11.87 (7.10) | 0.06 |

BMI, Body Mass Index; LEDD, Levodopa Equivalent Daily Dose; S&E, Modified Schwab and England Activities of Daily Living Scale; H&Y, Hoehn and Yahr; PIGD, Postural Instability and Gait Disorder; MDS-UPDRS, Movement Disorder Society Unified Parkinson’s Disease Rating Scale; MoCA, Montreal Cognitive Assessment; LFLT, Lexical Fluency Letter Test; MBNT, Modified Boston Naming Test, BJLO, Benton Judgment of Line Orientation; LNST, Letter Number Sequencing Test; SDMT, Symbol Digit Modalities Test; TMT, Trail Making Test; QUIP, Questionnaire for Impulsive-Compulsive Disorders in Parkinson’s Disease; GDS-15, Geriatric Depression Scale; STAI, State-Trait Anxiety Inventory; ESS, Epworth Sleepiness Scale; RBDSQ, REM Sleep Behavior Disorder Screening Questionnaire (values ≥ 5 are considered positive for RBD); SCOPA, Scales for Outcomes in Parkinson’s Disease – Autonomic; OH, orthostatic hypotension; NSD-ISS, Neuronal Synuclein Disease – Integrated Staging System. ON denotes the ON medication state. Values are *n* or mean (with corresponding % or SD), as appropriate, *: statistically significant difference between male and female, p<0.016 (BH threshold for FDR 0.05). m.p. = machine precision.

**eFigure 1: Sampling without Replacement for Cross-sectional Analysis**

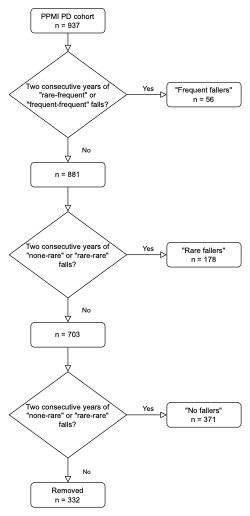

Of 937 unique PD patients in the PPMI, we conducted sampling without replacement for individuals with two consecutive years of available fall data. After sampling, 332 unique participants did not fulfill consecutive years’ definitions and were removed from the cross-sectional analysis.

**eFigure 2: Motor Differences between Never, Rare, And Frequent Fallers in PD**

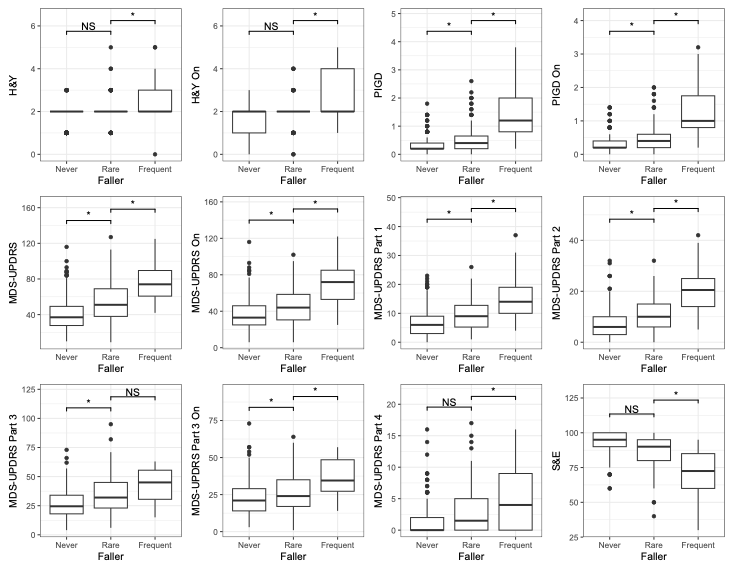

Boxplot displays the distribution of values across three groups. The central line represents the median, boxes indicate the interquartile range (IQR), and whiskers extend to 1.5 × IQR. Outliers are shown as individual points. Group differences were assessed using linear regressions adjusted for age, sex, and years since diagnosis. H&Y, Hoehn and Yahr; PIGD, Postural Instability and Gait Disorder; MDS-UPDRS, Movement Disorder Society Unified Parkinson’s Disease Rating Scale; S&E, Modified Schwab and England Activities of Daily Living Scale. ON denotes the ON medication state. *: statistically significant from rare, p<0.016 (BH threshold for FDR 0.05).

**eFigure 3: Non-Motor Differences Between Never, Rare, and Frequent Fallers in PD**

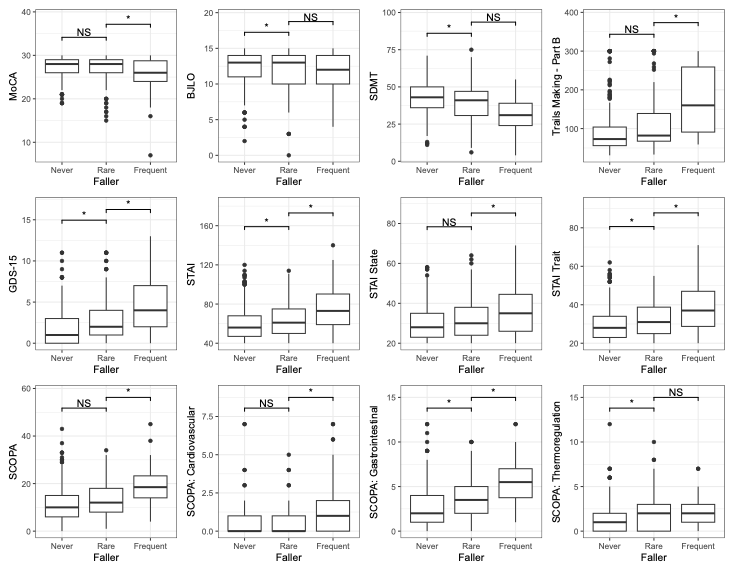

Boxplot displays the distribution of values across three groups. The central line represents the median, boxes indicate the interquartile range (IQR), and whiskers extend to 1.5 × IQR. Outliers are shown as individual points. Group differences were assessed using linear regressions adjusted for age, sex, and years since diagnosis. MoCA, Montreal Cognitive Assessment; BJLO, Benton Judgment of Line Orientation; SDMT, Symbol Digit Modalities Test; TMT, Trail Making Test, GDS-15, Geriatric Depression Scale; STAI, State-Trait Anxiety Inventory; SCOPA, Scales for Outcomes in Parkinson’s Disease – Autonomic. *: statistically significant from rare, p<0.016 (BH threshold for FDR 0.05).

**eFigure 4: Outcome Differences between Rare and Frequent Fallers in PD**

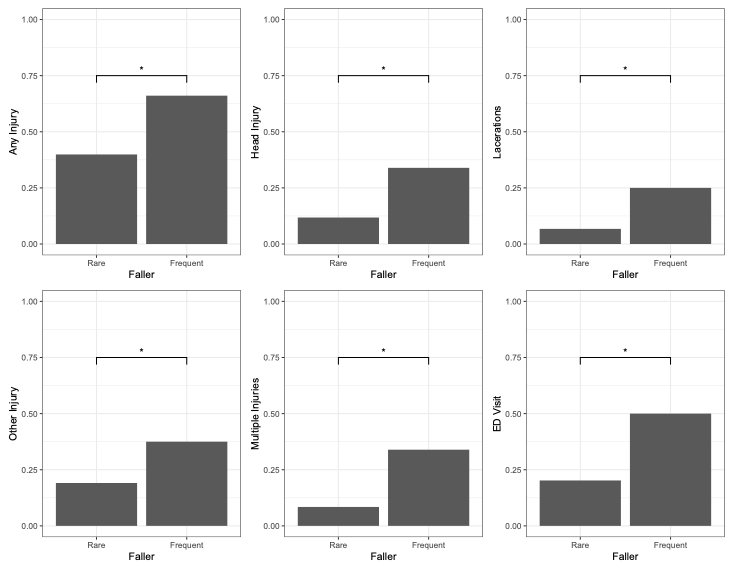

Bar plot shows the percentage of each outcome in each group. Group differences were assessed using logistic regressions adjusted for age, sex, and years since diagnosis. Frequent fallers showed significant differences from rare fallers for injuries, lacerations, multiple injuries, and ED visits. *: statistically significant from rare, p<0.016 (BH threshold for FDR 0.05).

**eFigure 5: Sex-based Differences in the Cross-Sectional PD Cohort**

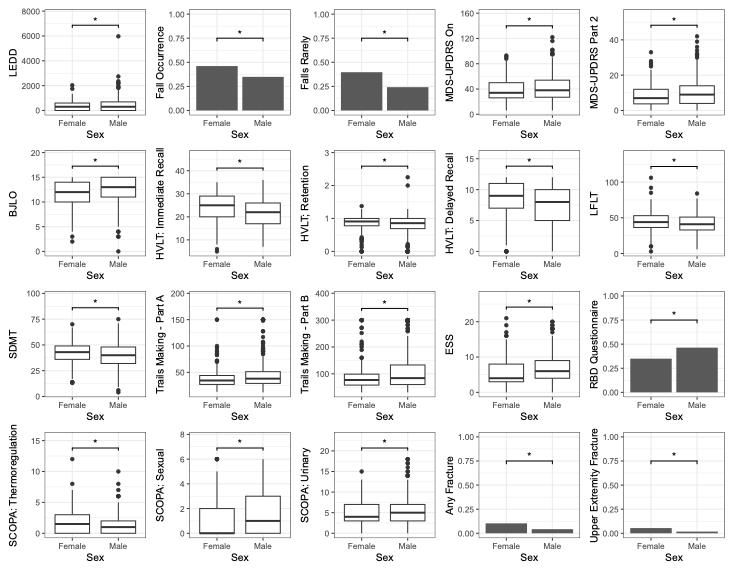

For continuous variables, boxplots display the distribution of values across two groups. The central line represents the median, boxes indicate the interquartile range (IQR), and whiskers extend to 1.5 × IQR. Outliers are shown as individual points. For categorical variables, bar plots show the percentage of each outcome in each group. Group differences were assessed using regressions adjusted for age, fall frequency, and years since diagnosis. Regressions were linear for continuous variables and logistic for categorical variables.

LEDD, Levodopa Equivalent Daily Dose; MDS-UPDRS, Movement Disorder Society Unified Parkinson’s Disease Rating Scale; BJLO, Benton Judgement of Line Orientation; HVLT, Hopkins Verbal Learning Test; LFLT, Lexical Fluency Letter Test; SDMT, Symbol Digit Modalities Test; TMT, Trail Making Test; ESS, Epworth Sleepiness Scale; RBQ, REM Behavioral Disorder; SCOPA, Scales for Outcomes in Parkinson’s Disease – Autonomic. *: statistically significant from rare, p<0.016 (BH threshold for FDR 0.05).

**PPMI STUDY TEAMS/CORES/COLLABORATORS FOR PUBLICATIONS**

Executive Steering Committee:

Kenneth Marek, MD1 (Principal Investigator); Caroline Tanner, MD, PhD9; Tanya Simuni, MD3; Andrew Siderowf, MD, MSCE12; Douglas

Galasko, MD27; Lana Chahine, MD41; Christopher Coffey, PhD4; Kalpana Merchant, PhD61; Kathleen Poston, MD40; Roseanne Dobkin,

PhD43; Tatiana Foroud, PhD15; Brit Mollenhauer, MD8; Dan Weintraub, MD12; Ethan Brown, MD9; Karl Kieburtz, MD, MPH23; Mark Frasier,

PhD6; Todd Sherer, PhD6; Sohini Chowdhury, MA6; Roy Alcalay, MD36 and Aleksandar Videnovic, MD47

Steering Committee:

Duygu Tosun-Turgut, PhD9; Werner Poewe, MD7; Susan Bressman, MD14; Jan Hammer15; Raymond James, RN22; Ekemini Riley,

PhD42; John Seibyl, MD1; Leslie Shaw, PhD12; David Standaert, MD, PhD18; Sneha Mantri, MD, MS62; Nabila Dahodwala, MD12;

Michael Schwarzschild47; Connie Marras45; Hubert Fernandez, MD25; Ira Shoulson, MD23; Helen Rowbotham2; Paola Casalin11

and Claudia Trenkwalder, MD8

Michael J. Fox Foundation (Sponsor): Todd Sherer, PhD; Sohini Chowdhury, MA; Mark Frasier, PhD; Jamie Eberling, PhD; Katie

Kopil, PhD; Alyssa O’Grady; Maggie McGuire Kuhl; Leslie Kirsch, EdD and Tawny Willson, MBS

Study Cores, Committees and Related Studies: (Include as applicable to the paper)

Project Management Core: Emily Flagg, BA1

Site Management Core: Tanya Simuni, MD3; Bridget McMahon, BS1

Strategy and Technical Operations: Craig Stanley, PhD1; Kim Fabrizio, BA1

Data Management Core: Dixie Ecklund, MBA, MSN4; Trevis Huff, BSE4

Screening Core: Tatiana Foroud, PhD15; Laura Heathers, BA15; Christopher Hobbick, BSCE15; Gena Antonopoulos, BSN15

Imaging Core: John Seibyl, MD1; Kathleen Poston, MD40

Statistics Core: Christopher Coffey, PhD4; Chelsea Caspell-Garcia, MS4; Michael Brumm, MS4

Bioinformatics Core: Arthur Toga, PhD10; Karen Crawford, MLIS10

Biorepository Core: Tatiana Foroud, PhD15; Jan Hamer, BS15

Biologics Review Committee: Brit Mollenhauer8; Doug Galasko27; Kalpana Merchant61

Genetics Core: Andrew Singleton, PhD13

Pathology Core: Tatiana Foroud, PhD15; Thomas Montine, MD, PhD40

Found: Caroline Tanner, MD PhD9

PPMI Online: Carlie Tanner, MD PhD9; Ethan Brown, MD9; Lana Chahine, MD41; Roseann Dobkin, PhD43; Monica Korell, MPH9

Site Investigators:

Charles Adler, PhD51; Roy Alcalay, MD36; Amy Amara, PhD52; Paolo Barone, PhD30; Bastiaan Bloem, PhD60 Susan Bressman,

MD14; Kathrin Brockmann, MD26; Norbert Brüggemann, MD59; Lana Chahine, MD41; Kelvin Chou, MD44; Nabila Dahodwala,

MD12; Alberto Espay, MD32; Stewart Factor, DO16; Hubert Fernandez, MD25; Michelle Fullard, MD52; Douglas Galasko, MD27;

Robert Hauser, MD19; Penelope Hogarth, MD17; Shu-Ching Hu, PhD21; Michele Hu, PhD58; Stuart Isaacson, MD31; Christine Klein,

MD59; Rejko Krueger, MD2; Mark Lew, MD49; Zoltan Mari, MD56; Connie Marras, PhD45; Maria Jose Martí, PhD34; Nikolaus

McFarland, PhD54; Tiago Mestre, PhD46; Brit Mollenhauer, MD8; Emile Moukheiber, MD28; Alastair Noyce, PhD63 Wolfgang

Oertel, PhD64; Njideka Okubadejo, MD65; Sarah O’Shea, MD39; Rajesh Pahwa, MD48; Nicola Pavese, PhD57; Werner Poewe, MD7;

Ron Postuma, MD55; Giulietta Riboldi, MD53; Lauren Ruffrage, MS18; Javier Ruiz Martinez, PhD35; David Russell, PhD1; Marie H

Saint-Hilaire, MD22; Neil Santos, BS51; Wesley Schlett47; Ruth Schneider, MD23; Holly Shill, MD50; David Shprecher, DO24; Tanya

Simuni, MD3; David Standaert, PhD18; Leonidas Stefanis, PhD38; Yen Tai, PhD29; Caroline Tanner, PhD9; Arjun Tarakad, MD20;

Eduardo Tolosa PhD34 and Aleksandar Videnovic, MD47

Coordinators:

Susan Ainscough, BA30; Courtney Blair, MA18; Erica Botting19; Isabella Chung, BS56; Kelly Clark24; Ioana Croitoru35; Kelly DeLano,

MS32; Iris Egner, PhD7; Fahrial Esha, BS53; May Eshel36; Frank Ferrari, BS44; Victoria Kate Foster57; Alicia Garrido, MD34; Madita

Grümmer59; Bethzaida Herrera50; Ella Hilt26; Chloe Huntzinger, BA52; Raymond James, BS22; Farah Kausar, PhD9; Christos Koros,

MD, PhD38; Yara Krasowski60; Dustin Le, BS17; Ying Liu, MD52; Taina M. Marques, PhD2; Helen Mejia Santana, MA39; Sherri

Mosovsky, MPH41; Jennifer Mule, BS25; Philip Ng, BS45; Lauren O’Brien48; Abiola Ogunleye, PGDip29; Oluwadamilola Ojo, MD65;

Obi Onyinanya, BS28; Lisbeth Pennente, BA31; Romina Perrotti55; Michael Pileggi, MS55; Ashwini Ramachandran, MSc12; Deborah

Raymond, MS14; Jamil Razzaque, MS58; Shawna Reddie, BA46; Kori Ribb, BSN,28; Kyle Rizer, BA54; Janelle Rodriguez, BS27;

Stephanie Roman, HS1; Clarissa Sanchez, MPH20; Cristina Simonet, PhD29; Anisha Singh, BS23; Elisabeth Sittig64; Barbara

Sommerfeld MSN16; Angela Stovall, BS44; Bobbie Stubbeman, BS32; Alejandra Valenzuela, BS49; Catherine Wandell, BS21; Diana

Willeke8; Karen Williams, BA3 and Dilinuer Wubuli, MB45

v. 15DEC2023Partners Scientific Advisory Board (Acknowledgement)

Funding: PPMI – a public-private partnership – is funded by the Michael J. Fox Foundation for Parkinson’s Research and

funding partners, including 4D Pharma, Abbvie, AcureX, Allergan, Amathus Therapeutics, Aligning Science Across Parkinson's,

AskBio, Avid Radiopharmaceuticals, BIAL, Biogen, Biohaven, BioLegend, BlueRock Therapeutics, Bristol-Myers Squibb, Calico

Labs, Celgene, Cerevel Therapeutics, Coave Therapeutics, DaCapo Brainscience, Denali, Edmond J. Safra Foundation, Eli Lilly,

Gain Therapeutics, GE HealthCare, Genentech, GSK, Golub Capital, Handl Therapeutics, Insitro, Janssen Neuroscience,

Lundbeck, Merck, Meso Scale Discovery, Mission Therapeutics, Neurocrine Biosciences, Pfizer, Piramal, Prevail Therapeutics,

Roche, Sanofi, Servier, Sun Pharma Advanced Research Company, Takeda, Teva, UCB, Vanqua Bio, Verily, Voyager

Therapeutics, the Weston Family Foundation and Yumanity Therapeutics.

1 Institute for Neurodegenerative Disorders, New Haven, CT

2 University of Luxembourg, Luxembourg

3 Northwestern University, Chicago, IL

4 University of Iowa, Iowa City, IA

5 VectivBio AG

6 The Michael J. Fox Foundation for Parkinson’s Research, New York, NY

7 Innsbruck Medical University, Innsbruck, Austria

8 Paracelsus-Elena Klinik, Kassel, Germany

9 University of California, San Francisco, CA

10 Laboratory of Neuroimaging (LONI), University of Southern California

11 BioRep, Milan, Italy

12 University of Pennsylvania, Philadelphia, PA

13 National Institute on Aging, NIH, Bethesda, MD

14 Mount Sinai Beth Israel, New York, NY

15 Indiana University, Indianapolis, IN

16 Emory University of Medicine, Atlanta, GA

17 Oregon Health and Science University, Portland, OR

18 University of Alabama at Birmingham, Birmingham, AL

19 University of South Florida, Tampa, FL

20 Baylor College of Medicine, Houston, TX

21 University of Washington, Seattle, WA

22 Boston University, Boston, MA

23 University of Rochester, Rochester, NY

24 Banner Research Institute, Sun City, AZ

25 Cleveland Clinic, Cleveland, OH

26 University of Tübingen, Tübingen, Germany

27 University of California, San Diego, CA

28 Johns Hopkins University, Baltimore, MD

29 Imperial College of London, London, UK

30 University of Salerno, Salerno, Italy

31 Parkinson’s Disease and Movement Disorders Center, Boca Raton, FL

32 University of Cincinnati, Cincinnati, OH

34 Hospital Clinic of Barcelona, Barcelona, Spain

35 Hospital Universitario Donostia, San Sebastian, Spain

36 Tel Aviv Sourasky Medical Center, Tel Aviv, Israel

37 St. Olav’s University Hospital, Trondheim, Norway

38 National and Kapodistrian University of Athens, Athens, Greece

39 Columbia University Irving Medical Center, New York, NY

40 Stanford University, Stanford, CA

41 University of Pittsburgh, Pittsburgh, PA

42 Center for Strategy Philanthropy at Milken Institute, Washington D.C.

43 12, New Brunswick, NJ

44 University of Michigan, Ann Arbor, MI

45 Toronto Western Hospital, Toronto, Canada

46 The Ottawa Hospital, Ottawa, Canada

47 Massachusetts General Hospital, Boston, MA

48 University of Kansas Medical Center, Kansas City, KS

49 University of Southern California, Los Angeles, CA

50 Barrow Neurological Institute, Phoenix, AZ

v. 15DEC202351 Mayo Clinic Arizona, Scottsdale, AZ

52 University of Colorado, Aurora, CO

53 NYU Langone Medical Center, New York, NY

54 University of Florida, Gainesville, FL

55 Montreal Neurological Institute and Hospital/McGill, Montreal, QC, Canada

56 Cleveland Clinic-Las Vegas Lou Ruvo Center for Brain Health, Las Vegas, NV

57 Clinical Ageing Research Unit, Newcastle, UK

58 John Radcliffe Hospital Oxford and Oxford University, Oxford, UK

59 Universität Lübeck, Luebeck, Germany

60 Radboud University, Nijmegen, Netherlands

61 TransThera Consulting

62 Duke University, Durham, NC

63 Wolfson Institute of Population Health, Queen Mary University of London, UK

64 Philipps-University Marburg, Germany

65 University of Lagos, Nigeria

v. 15DEC2023
